# Automatic Electrocardiogram Detection of Suspected Hypertrophic Cardiomyopathy: Application to Wearable Heart Monitors

**DOI:** 10.1101/2021.05.21.21257601

**Authors:** Max Denis, Mulatu Bachoro, Winta Gebreslassie, Timothy Oladunni

## Abstract

In this work, an automatic detection algorithm for hypertrophic cardiomyopathy (HCM) is presented. Of particular interest is the algorithm’s ability to differentiate HCM subjects and healthy volunteers from a single lead ECG dataset. Suspected HCM subjects are identified by the primary clinical abnormality associated with HCM, left ventricular hypertrophy (LVH). In total, *n*=43 human subjects ECG datasets are investigated: *n*=21 healthy volunteers and *n*=22 left ventricular hypertrophy (LVH) patients. Significant differences of *p*-value 0.01 and 0.04 were found for the respective ECG parameters, S-wave amplitude and ST-segment, when differentiating between the LVH patients and healthy human volunteers.

## INTRODUCTION

Hypertrophic cardiomyopathy is the most common condition responsible for sudden cardiac death (SCD) in young athletes [1-3] with an estimated annual mortality of 1-2 %. In a large case series of SCD in 1,866 young athletes, HCM was identified in nearly 40 % of these cases [3]. Studies have also revealed a strong preponderance for SCD in African-American (AA) athletes who compete in sports with sudden movements and adrenergic surges such as football or basketball [4]. Unfortunately, over 80 % of affected individuals are asymptomatic before SCD, which often occurs during exercise or in its aftermath. With a heightened awareness of SCD in young athletes, screening methods have been developed to try and prevent these events from occurring.

Previous studies have attempted to assess the electrophysiological signature of HCM by visually inspecting the standard 12-lead electrocardiogram (ECG). ECG signal originates from the electrical activity of the heart that coordinates the contraction and relaxation of the different chambers of the heart. The analysis of ECG signals and detection of its characteristic points can be used to identify various heart rhythm abnormalities, chest pains and other diseases. One cardiac cycle of an ECG signal comprises of the P-wave, T-wave, and QRS complex (comprised of Q-wave, R-wave, and S-wave). ECG signals of HCM have reported abnormal signatures, such as abnormal Q waves, wide and high amplitude QRS complexes, ST-segment displacement as well as giant inverted T-waves, yet ECG biomarkers are not used for risk stratification. The American Heart Association currently employs history and physical examination alone during the pre-participation physical exam (PPE), which clears a young athlete for participation in sports. In the United States, the American Heart Association (AHA) consensus expert panel does not endorse mandatory athlete screening with the inclusion of ECG.

In the absence of reliable ECG biomarkers for predicting SCD in HCM from visual inspection, more sophisticated approaches are required. Real-time automatic ECG detection methods on a wearable mobile heart monitor can provide automatic detection of ECG features and continuous monitoring for risk-stratifications of HCM. Rahman *et al*. [5], presented a patient classifier to automatically detect patients affected by HCM based on the standard 12-lead ECG. They classified a patient as HCM, if the majority of the beats show HCM beat morphology. Two hundred and sixty-four standard ECG features, such as time intervals and waveforms amplitude, were extracted by feature selection, and used to perform machine learning classification reaching a precision of 84% (0.89 sensitivity, 0.93 specificity).

In this paper, we will present an automatic ECG feature detection method on data collected from single lead wearable heart monitor. The goal is to identify the key ECG parameters differentiating healthy individuals and those at-risk for HCM outside of a clinical setting.

## LEFT VENTRICULAR HYPERTROPHY

The chief abnormality associated with HCM is left ventricular hypertrophy (LVH) [6]. LVH is a left ventricular diastolic dysfunction resulting from impaired relaxation and filling of the stiff and hypertrophied left ventricle. The degree and distribution of LVH is variable: mild hypertrophy or extreme myocardial thickening may be seen.

LVH results in the following ECG features: increased R-wave amplitude in the left-sided ECG leads (I, aVL and V4-6) and increased S-wave depth in the right-sided leads (III, aVR, V1-3)[5, 6]. The thickened left ventricle wall leads to prolonged depolarization (increased R wave peak time > 50 ms in leads V5 or V6) and delayed repolarization (ST-segment depression and T-wave inversion) in the lateral leads.

## MATERIALS AND METHODS

### Data Description

Here, we describe the ECG datasets for healthy human volunteers during physical activity, and LVH patients at rest. Chest electrocardiography (ECG) datasets of the healthy human volunteers were obtained from two databases. First, *n*=9 single lead chest ECG signals of healthy human volunteers were obtained from the PhysioNet database recorded at a sampling frequency of 256 Hz [7, 8]. The ECG signals were recorded in four different exercise conditions:

- While walking on a treadmill.
- While running on a treadmill.
- While using an exercise bike set to a low resistance (giving high cycling speeds).
- While using an exercise bike set to a high resistance (giving low cycling speeds).

Second, single lead chest ECG signals of *n*=12 healthy human volunteers, recorded at a sampling frequency of 125 Hz, were obtained from the 2015 IEEE Signal Processing Cup database [9]. During data recording, each subject ran on a treadmill with changing speeds of 6-15km/h for one minute.

The LVH patient ECG dataset is a subset of The Massachusetts General Hospital/Marquette Foundation (MGH/MF) Waveform Database [10]. The MGH/MF database is a comprehensive collection of electronic recordings of hemodynamic and electrocardiographic waveforms of stable and unstable patients in critical care units, operating rooms, and cardiac catheterization laboratories. In total, *n*=22 LVH three lead ECG recordings, with a sampling frequency of 360Hz.

In normal healthy people, the expected ECG changes from at rest to during exercise are the following: RR interval decreases, QRS complex experiences minimal shortening, ST segment becomes upsloping, and QT interval experiences a rate-related shortening [11]. Meanwhile, LVH patients’ measured ECGs at rest, relative to normal ECGs, display the following characteristics: 1) a high R-wave peak time (> 50 ms) which is associated with the widening of the QRS complex, and is the opposite effect when normal healthy people exercise; and 2) ST segment depression occurs, which is opposite to the observed ST upsloping when normal healthy people exercise. Therefore, we expect the ECG parameters of the two cohorts to be different due to the contrasting effects on the heart during exercise and having LVH at rest.

### Proposed Detection Algorithm

The basic strategy behind the ECG feature detection algorithm is to first detect the heartbeat, namely the R-to-R wave interval. Furthermore, calculate the QRS complex using a template with the detected ECG R-wave peak positions, thereafter detect the P and T waves. Prior to the R-wave detection the ECG signals are preprocessed. Fig. 1 shows the ECG preprocessing procedure. To remove the 50 Hz interference, a notch filter (at 50 Hz) is used. To suppress the baseline wander, a two-order smooth filter (0.6 second window) is applied to ECG. Finally, a bandpass FIR filter between 0.5–80 Hz is used to further suppress the noise of the ECG.

**Fig. 1.**
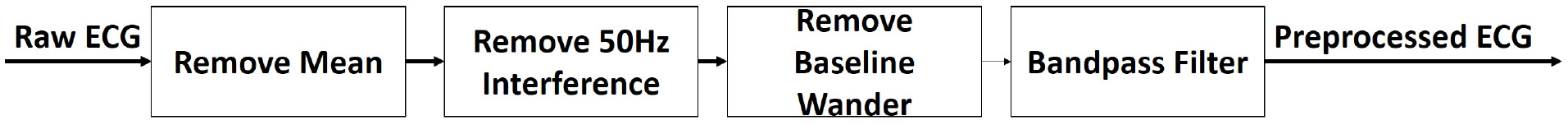
ECG preprocessing procedure (preprocessing-ECG).

ECG signal can be intermixed with many kinds of unwanted noises including body motion. The quality of the ECG signal has been associated with the problem of false alarms. Automatic quality detection assessment and classification of ECG signals can play a vital role in the development of real-time ECG diagnosis. Li *et al*. [12] proposed the signal quality index “bSQI” makes a comparison of two beat detectors on a single ECG lead. The two detectors where ‘ep_limited’ [13] and ‘wqrs’ [14] The bSQI ranges between 0 and 1 for the kth beat is defined as:

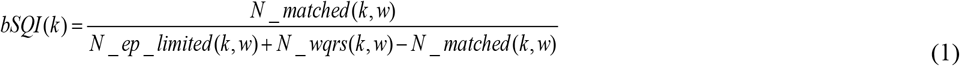

where *N* _ *matched* (*k, w*) is the beat number agreed upon (within *γ* = 150 *ms*), *N* _ *ep* _ *limited* (*k, w*) is the beat number detected by ‘*ep_limited*’ and *N wqrst(k, w)* is the beat number detected by ‘*wqrs*’.

The ECGPUWAVE algorithm is used to detect the QRS complexes and locating the beginning, peak, and end of the P, QRS, and ST-T waveforms [8]. The QRS detector is based on the algorithm of Pan and Tompkins using the slope information [15, 16]. The signal slope in the decision rule any possible detection should have a maximum slope within of that of the previous QRS complexes. To detect the T wave, a search window is defined as a function of the heart rate. The algorithm determines the type of T wave (regular, inverted, biphasic +-, or biphasic -+). ECGPUWAVE classifies each T-wave as type 0 (normal), 1 (inverted), 2 (positive monophasic), 3 (negative monophasic), 4 (biphasic negative-positive), or 5 (biphasic positive-negative).

### Data Analysis

Test for significant differences in the ECG parameters (QtT interval, ST-segment, S-wave amplitude, and R wave peak time) between healthy volunteers and LVH patients. Significant tests were conducted with Tukey’s HSD *post-hoc* tests, *p*-values < 0.05 were considered to indicate statistical significance. To assess the statistical performance of the ECG parameters that best delineates healthy human subjects and LVH patients a receiver operating characteristic (ROC) analysis is performed. An optimal cutoff value, which maximizes the sensitivity and specificity of the ROC curve, was established.

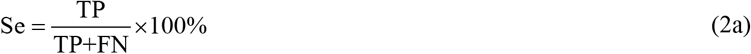

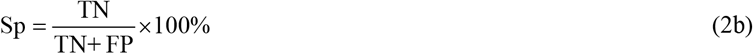

where Se is sensitivity, Sp is specificity, TP is true positive, TN is true negative, FP is false positive, and FN is false negative. Statistical analyses were performed using MATLAB software (The MathWorks, Natick, MA, USA).

## RESULTS

Fig. 2 demonstrates the utility of the automatic detection algorithm of a 10 second ECG for a heathy subject and LVH patients. Fig. 2a is an ECG of healthy subject running on a treadmill. Fig. 2b demonstrates the automatic detection results of the ECGPUWAVE for the QRS complex, T_Onset_ (beginning of the T-wave), T_Peak_ (peak amplitude of the T-wave), and T_Offset_ (end of the T-wave) features of the ECG signal in Fig. 2a.

**Fig. 2.**
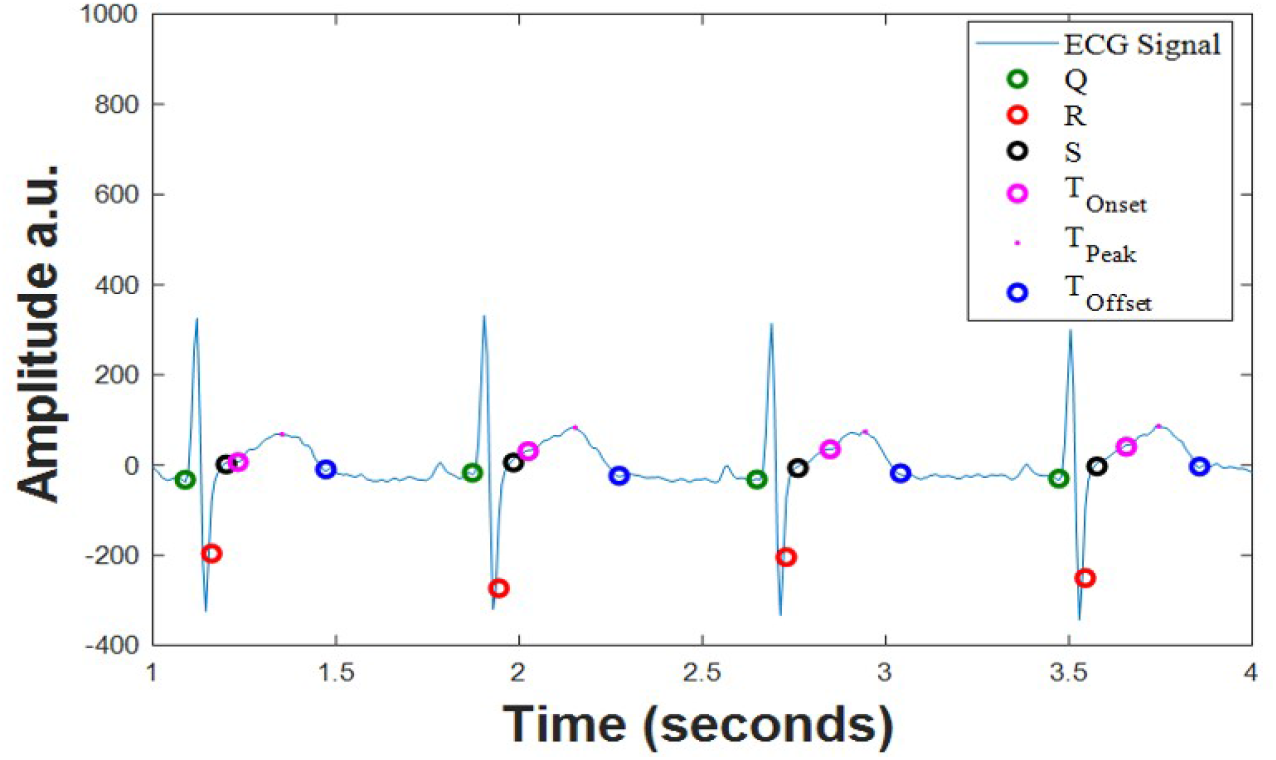
LVH patient processed ECG signal with automatically detected ECG features: QRS complex, T_Onset_, T_Peak_, and T_Offset_.”

Fig. 3 shows the comparative boxplots of the ECG parameters (QT interval, ST-segment, T-wave inversion, and R peak time) for the healthy subjects and the LVH patients. Fig. 3a illustrates that the median QT interval (red line through the box) for the LVH patients (QT interval= 0.44s) is similar to the healthy subjects (QT interval= 0.48s). Fig. 3b shows a significant separation between the median ST-segment values for the LVH patients (ST-segment= -0.0011) than healthy human subjects (ST-segment= 0.47s). Similarly, Fig. 3(c-d) shows significant separation between LVH patients and healthy volunteers in the median values of the S-wave amplitude and R-wave to peak time.

**Fig. 3.**
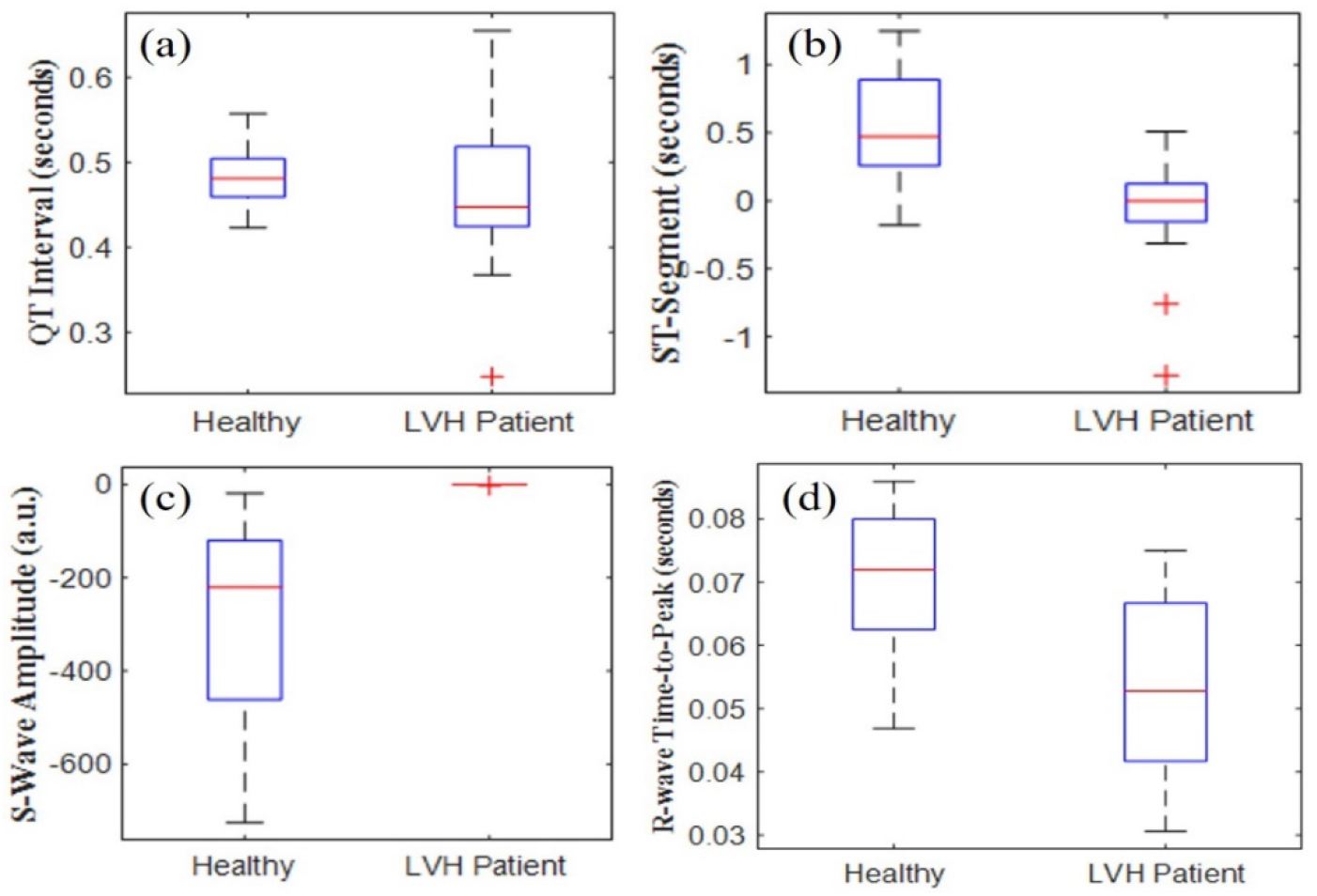
Boxplot: (a) QT interval (b) ST segment, (c) S-wave amplitude, and (d) R-wave peak.

The post hoc test results are summarized in Table 1. The results reveal significant differences between healthy subjects walking or running on a treadmill and LVH patients using ST segment, with p values less than 0.04. Other notable results from Table 1, is the p-value of 0.01 for significant differences in S-wave amplitude for LVH patients and healthy subjects on the treadmill.

**Table 1.** Statistical difference (*p*-value) for levels of activity.

| ECG Parameters |  | QT | ST | S Amp. | R Peak |
| --- | --- | --- | --- | --- | --- |
| LVH | Biking | 0.69 | 0.04 | 0.08 | 0.26 |
|  | Treadmill /Walking | 0.92 | 0.03 | 0.01 | 0.11 |
| Biking | Treadmill /Walking | 0.45 | 0.55 | 0.69 | 0.77 |
|  | LVH | 0.69 | 0.04 | 0.08 | 0.26 |
| Treadmill /Walking | Biking | 0.45 | 0.55 | 0.69 | 0.77 |
|  | LVH | 0.92 | 0.03 | 0.01 | 0.11 |

QT: QT interval, ST: ST segment, S Amp.: S-wave Amplitude, and R Peak: R-wave peak time.

Fig. 4 shows the ROC curve between LVH patients and healthy subjects for QT, ST-segment, S-wave amplitude and R-wave time to peak. From observation, ST-segment and S-wave amplitude demonstrating high sensitivity for a range of specificities.

**Fig. 4.**
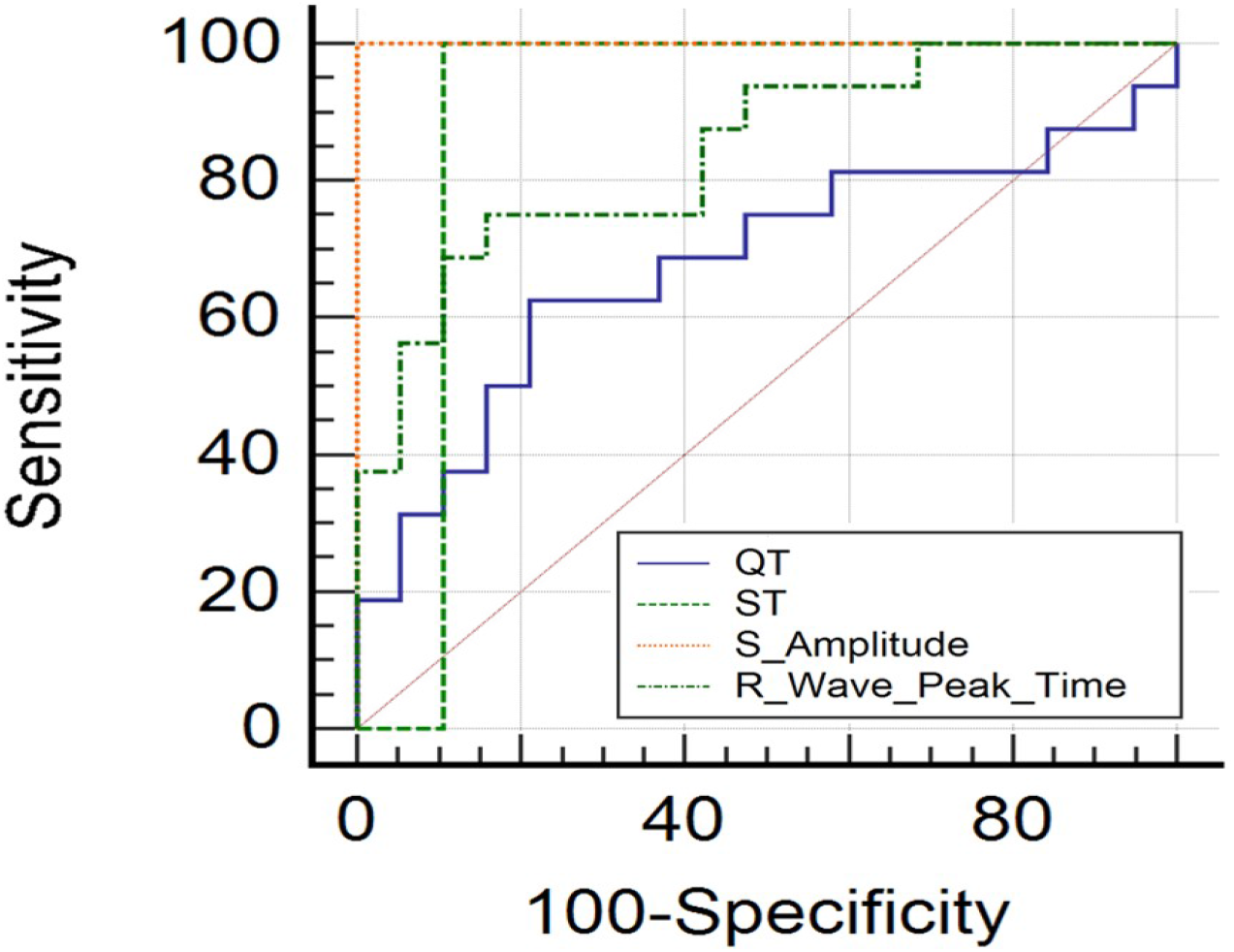
ROC curve

According to the ROC analysis, the optimal cut-off values for the ECG parameters QT interval, ST-segment, S-wave amplitude, and R-wave peak time are 0.45s, 0.51s, 19 a.u., and 0.51s, respectively.

The sensitivity (Se), specificity (Sp) and area under the curve (AUC) results after applying the cut-off values are summarized in Table 2. The ST-segment and S-wave amplitude demonstrate the best results.

**Table 2.** ROC analysis of ECG parameters.

| ECG Parameters | Se (%) | Sp (%) | AUC (%) |
| --- | --- | --- | --- |
| QT Interval<br>Cutoff $\leq 0.45s$ | 63 | 79 | 67 |
| ST Segment<br>Cutoff $\leq 0.51s$ | 100 | 90 | 89 |
| S-wave Amplitude<br>Cutoff $\leq 19$ a.u. | 100 | 100 | 100 |
| R-wave Peak Time Cutoff $\leq 0.51s$ | 75 | 84 | 84 |

## DISCUSSION

The 12-lead ECG is a fundamental initial diagnostic modality for the early evaluation of a patient suspected of having HCM. Previous studies [17-19] of patients with mutations in cardiac myofilaments, ECG had an accuracy of sensitivity ∼60% and specificity∼98% in the diagnosis of HCM as echocardiography (left ventricular wall thickness: 13 mm). These results are similar to our QT and R-wave time to peak analysis. However, access to 12-lead ECGs is not always readily available away from a clinical setting. Thus, wearable heart monitors, with automatic ECG feature detection algorithms, have been utilized for remotely monitor suspected patients with heart ailments. Additionally, these heart monitors have been integrated into commercial mobile personal devices, making their availability ubiquitous.

The principal finding of the current study is the following: abnormal ST-segment, and S-wave amplitude were the best predictors for differentiating healthy subjects and LVH patients, specifically for ST-segment < 0.51 seconds and S-wave amplitude < 19 a.u. Other observations include the overlap in boxplot analysis of QT interval and R-wave time-to-peak. This may be evidence of a HCM phenotype, where LVH patients with HCM have a normal ECG. McLeod *et al*. [20] compared HCM patients with normal and abnormal ECGs, and found that those with a normal ECG presented at an older age and with less severe disease expression. However, our study is limited in the small number of healthy subjects and LVH patient datasets, as well as their asynchronous comparison of ECG measurements during physical activity and at rest, respectively. Additionally, only 4 ECG parameters were presented. Future works will include larger datasets and additional ECG parameters.

## CONCLUSION

The utility of wearable ECG monitors for heart diagnosis was presented. The results of this study demonstrate the feasibility of a single lead wearable heart monitor to automatically detect ECG features from datasets of healthy subjects and LVH patients. Additionally, ECG parameters ST segment and S-wave amplitude successfully differentiated LVH patients and healthy subjects. These results parallel the ECG characteristics of HCM including the presence of LVH ECG features and ST segment abnormalities. However, due to our study limitations further investigation is needed.

## Data Availability

All datasets used in this manuscript are publicly available.

## ACKNOWLEDGMENT

This material is based upon work supported by the National Science Foundation under Grant No. 2032345. Data Availability Statement: All datasets used in this manuscript are publicly available.

## REFERENCES

1. Pelliccia, A., et al., The upper limit of physiologic cardiac hypertrophy in highly trained elite athletes. New England Journal of Medicine, 1991. 324(5): p. 295–301.

2. Basavarajaiah, S., et al., Prevalence of hypertrophic cardiomyopathy in highly trained athletes: relevance to pre-participation screening. Journal of the American College of Cardiology, 2008. 51(10): p. 1033–1039.

3. Maron, B.J., et al., Sudden deaths in young competitive athletes. Circulation, 2009. 119(8): p. 1085–1092.

4. Harmon, K.G., et al., Incidence of sudden cardiac death in National Collegiate Athletic Association athletes. Circulation, 2011. 123(15): p. 1594–1600.

5. Goldberger, A.L., Left ventricular hypertrophy: Clinical findings and ECG diagnosis.

6. Harrigan, R.A. and K. Jones, ABC of clinical electrocardiography: Conditions.

7. Jarchi, D. and A.J. Casson, Description of a database containing wrist PPG signals recorded during physical exercise with both accelerometer and gyroscope measures of motion. Data, 2017. 2(1): p. 1.

8. Goldberger, A., et al., Components of a new research resource for complex physiologic signals. PhysioBank, PhysioToolkit, and Physionet, 2000.

9. Zhang, Z., Z. Pi, and B. Liu, TROIKA: A general framework for heart rate monitoring using wrist-type photoplethysmographic signals during intensive physical exercise. IEEE Transactions on biomedical engineering, 2014. 62(2): p. 522–531.

10. Welch, J., et al., The Massachusetts General Hospital-Marquette Foundation hemodynamic and electrocardiographic database–comprehensive collection of critical care waveforms. Clinical Monitoring, 1991. 7(1): p. 96–97.

11. Sharma, S., Practical ECG for exercise science and sports medicine. 2010: Human Kinetics 1.

12. Li, Q., R.G. Mark, and G.D. Clifford, Robust heart rate estimation from multiple asynchronous noisy sources using signal quality indices and a Kalman filter. Physiological measurement, 2007. 29(1): p. 15.

13. Hamilton, P.S. and W.J. Tompkins, Quantitative investigation of QRS detection rules using the MIT/BIH arrhythmia database. IEEE transactions on biomedical engineering, 1986(12):p. 1157–1165.

14. Zong, W., G. Moody, and D. Jiang. A robust open-source algorithm to detect onset and duration of QRS complexes. in Computers in Cardiology, 2003. 2003. IEEE.

15. Pan, J. and W.J. Tompkins, A real-time QRS detection algorithm. IEEE transactions on biomedical engineering, 1985(3):p. 230–236.

16. Laguna, P., R. Jané, and P. Caminal, Automatic detection of wave boundaries in multilead ECG signals: Validation with the CSE database. Computers and biomedical research, 1994. 27(1): p. 45–60.

17. Wigle, E.D., et al., Hypertrophic cardiomyopathy. The importance of the site and the extent of hypertrophy. A review. Progress in cardiovascular diseases, 1985. 28(1): p. 1–83.

18. Wigle, E.D., et al., Hypertrophic cardiomyopathy: clinical spectrum and treatment. Circulation, 1995. 92(7): p. 1680–1692.

19. Charron, P., et al., Diagnostic value of electrocardiography and echocardiography for familial hypertrophic cardiomyopathy in a genotyped adult population. Circulation, 1997. 96(1): p. 214–219.

20. McLeod, C.J., et al., Outcome of patients with hypertrophic cardiomyopathy and a normal electrocardiogram. Journal of the american College of Cardiology, 2009. 54(3): p. 229–233.

